# Side Effects and Perceptions Following Sinopharm COVID-19 Vaccination

**DOI:** 10.1101/2021.06.28.21258847

**Authors:** Balsam Qubais Saeed, Rula Al-Shahrabi, Shaikha Salah Alhaj, Zainab Mansour Alkokhardi, Ahmed Omar Adrees

## Abstract

**Background:** Vaccines are one of the best interventions developed for eradicating COVID-19, the rapid creation of vaccinations was increased the risk of vaccine safety problems. The aim of this study to provide evidence on Sinopharm COVID-19 vaccine side effects which is approved by the United Arab Emirates (UAE).

**Methods:** A cross-sectional survey study was conducted between January and April 2021 to collect data on the effects of COVID-19 vaccine among individuals in the UAE. Demographic data, chronic conditions, side effects of the 1^st^ and 2^nd^ dose toward the vaccination, and the response of unwilling taking COVID-19 vaccine were reported.

**Results:** The most common side effects of post 1^st^ dose vaccination among participants (≤49 years old vs >49 years) were normal injection site pain 42.2%, fatigue 12.2%, and headache 9.6%, while pain at the vaccination site 32.6%, fatigue16.3%, lethargy13.7%, headache10%, and tenderness 10% were the most side effects of the post 2^nd^ dose of vaccination in both groups. All the side effects in both doses were more prevalent among the participants ≤ 49-year-old group.

Among two groups (females vs males), the study revealed the increase in the number of females that suffered from the vaccine side effects compared with males in both doses. The most prevalence adverse reactions of first dose in (female’s vs males) were fatigue (15.8% vs 3.75%), lethargy (12.6% vs 1.25%), headache (10.5% vs 7.5%), while in 2^nd^ dose were fatigue (20% vs 7.5%), sever injection site pain (10.5% vs 2.5%). The most common reason of not willing to take the COVID-19 vaccine among the participants were the vaccines are not effective, and the participants were not authorized to take vaccine.

**Conclusion:** The 1^st^ and 2^nd^ dose post-vaccination side effects were mild, predictable, and there were no hospitalization cases, this data will help to reduce the vaccine hesitancy.

## Introduction

The severe acute respiratory syndrome coronavirus-2 (SARS-CoV-2) causing coronavirus disease 2019 (COVID-19) [1]. The virus has spread fast over the world, resulting various levels of illnesses. On March 11, 2020, was announced that SARS-CoV-2 is a worldwide pandemic that has lasted through this moment [2].

Even though numerous therapeutic medications have been presented to resist COVID-19, they remain supportive and still require more randomized control studies to determine their efficacy and potency. [3, 4]. develop the vaccines is perhaps the best way to stopping this pandemic.

Vaccines are one of the best interventions developed for eradicating COVID-19, it saving millions lives annually, moreover, the best option remains an effective, safe vaccine, and without serious adverse reactions. The lack of effective and approved treatment of COVID-19 has triggered a vaccine development race, with 259 COVID-19 vaccine projects underway from November 11, 2020. The rapid creation of vaccinations was increased the risk of vaccine safety problems [5, 6].

Several candidate COVID-19 vaccines were developed from diverse platforms, one of them BBIBP-CorV vaccine (also known as the Sinopharm COVID-19 vaccine) that was made by Chinese state-owned pharmaceutical business called Sinopharm in China that adopted by the UAE [7].

Sinopharm COVID-19 vaccine is an inactivated vaccine that introduces a dead copy of SARS-CoV-2 into the body by a two-dose schedule, with 14 or 21 days between the 2 doses. However, by inserting the vaccine dose intramuscularly, the dead antigens from the virus are employed to make antibodies that prepare the immune system for future attacks by the virus. [8].

The traditional inactivated whole-virus vaccines don’t lead to a clinical disease. In this technology the inactivated viruses maintain their ability to replicate in vivo with mild or no symptoms. [9]. Clinical trial 1 and 2 of Sinopharm COVID-19 vaccine were done in China over 1 trial for each, with a total enrolment of 640 persons showed that the vaccine triggered a COVID-19 neutralizing antibody response with a low rate of adverse reactions. The most common side effects were fever and pain at the site of injection and fever, but were mild, self-limiting, and didn’t required treatment [10].

Phase 3 was done over 4 trials in the following countries, the UAE, Bahrain, Egypt, Jordan, Peru, and Argentina, with a total of 69,000 people enrolled. The UAE approved the vaccine developed by the Chinese state-owned Sinopharm on December 9, 2020, UAE announcements that the vaccine was 86% efficacious, according to interim results of its phase 3 trial [8].

Had administered over 2 million doses of the vaccine as of mid-January, the UAE Ministry of Health also reported the vaccine to be 100% effective in the prevention of mild and severe COVID-19 cases [11, 12, 13, 14], Moreover, WHO Announced that the most side effects of Sinopharm COVID-19 vaccine in three clinical trials on 16,671 participants aged 18–59 years were mild to moderate in addition, headaches, fatigue, and injection site reactions were the most common side effects of this vaccine. [15].

Published data to support adverse reaction of Sinopharm COVID-19 vaccine are lacking, two studies only focus on Sinopharm COVID-19vaccine [16, 17], moreover, fear of the new vaccine is a driver of vaccine hesitancy.

The knowledge about what happens post vaccination in the actual world among the general population is still modest, Thus, by describing what to expect after 1^st^ and 2^nd^ dose of vaccination will help in lowering the apprehension about this type vaccines, increased the public confidence in the vaccines, safety, and accelerates the vaccination process against COVID-19. [16, 17]. The results of this study will be reassuring to those who are fearful of the Sinopharm COVID-19 vaccine. So, the goal of this study to provide evidence on Sinopharm COVID-19 vaccine side effects after receiving 1st and 2nd dose of it, which is approved by UAE.

## Methodology

### Study Design & Participants

This cross-sectional survey-based study was carried out from 10^th^ January 2021 to 30^th^ April 2021, to evaluate the Sinopharm COVID-19 vaccine adverse reactions among residents in the UAE. The study utilized a self-administered online survey created on Google Forms of platform which had been randomly delivered to adults’ individuals (18 years and older) using social media sites (Facebook, Email, and WhatsApp). Potential participants were directed to a page that included brief introduction to the aim and purpose of the study in addition to instructions on how to complete the survey. Informed consent form that included statements about voluntary participation and anonymity was sought from all the respondents prior to data collection by sending a standardized general invitation letter with the survey link to accept or decline to participate in the study. The participant who declined consent were not permitted to open the survey and participate in the study, and participants could withdraw from the survey at any time in line with stipulations of the World Medical Association Declaration of Helsinki Ethical principles [18]. The members who clicked on the link were directed to the Google forms and to avoid the missing data, the participants were requested to fill all the questions of the survey or else could not proceed to the next section. No incentives or compensations have been given to participants. The total number of respondents was calculated.

Out of 1102 survey received from respondents, 1080 participants were included in this study aged 18 years and above from different emirates and nationalities. The study sample included the participants who are either vaccinated with the first dose or the second dose of Sinopharm COVID-19 vaccine. Other participants were didn’t get any COVID-19 vaccine during the early peak of the vaccination campaign in UAE.

The study was approved by the Research Ethics Committee of university of Sharjah in the on: 22/3/2021, with the reference No: REC-21-02-09-07.

### Survey & Data collection

The Survey was designed based on previous extensive literature search studies, and guidelines of WHO, MOHAP, DOH in UAE on the expected Adverse reactions post Sinopharm COVID-19 vaccine. the questions of survey were multiple choice, the survey was created in English and Arabic languages. [8, 15, 19], it was validated by a group of experts who provided feedback on the different items of the survey.

The survey were included three sections, the first section included 7 demographic questions such as (gender, age, marital status, education level, employment, nationality, and, the emirate of living) second section reviewed participant’s chronic conditions such as (acute cancer, autoimmune diseases, chronic respiratory diseases, diabetes, hypertension, obesity, heart disease, severe allergic reactions to vaccination, receiving immunotherapy or inhibitor therapy, severe anemia, liver diseases, and no chronic condition). The last section was related to Sinopharm COVID-19 vaccine side effects, in the first order participant has been asked about his/her COVID-19 vaccination status to determine the scope of the rest of the questions, however, if the participant received the vaccine questions were as following (number of doses administered and the previous infection to COVID-19), then questions regarding vaccine 1st and 2nd dose side effects were asked separately based on each dose side effects, mentioning : (normal/severe pain at the vaccination site, tenderness, redness, pruritus at the vaccination site, fever, headache, fatigue, nausea, diarrhea, cough, allergy, muscle pain, abdominal pain, back pain, lethargy).

If the participant did not receive the any COVID-19 vaccine, a question to clarify the reasons behind the participant’s hesitancy takes place. For pilot testing, a questionnaire was passed randomly to 15 participants recently vaccinated and filled the questionnaire after taking the two doses and have been excluded from the study. The Cronbach’s alpha test of internal consistency was used to evaluate survey reliability. The overall reliability was equal to 0.81, which indicates that the survey tool was reliable with good internal consistency [20].

### Statistical analysis

The Statistical Package for the Social Sciences (SPSS) version 22.0 (SPSS Inc. Chicago, IL, USA, 2013) was used to carry out descriptive statistics of 1080 participants for the demographic variables (gender, age, education level, marital status, employment status, nationality and Emirate of residence. In addition, health and chronic condition, COVID-19-related anamnesis such as (vaccination status, previous infection, number of doses) and the reasons behind participants vaccination hesitancy. All data have been presented in both frequencies and percentages.

In our study the mean age was 37.2±13.1, given the fact that age has been classified into four groups ranged from the minimum of 18 to the maximum of 80 years old, the participants of 49 years old selected to be a cutting point to evaluate the differences of participants side effects between two groups (≤49 vs >49) using a chi-squared test. Similarly, chi-squared test was performed to assess the correlation between the presence of vaccine side effects and gender tolerability.

## Results

### Demographic characteristics

Table 1. shows the demographic data of participants, 760 (70.4%) were females, 320 (29.6%) were males. the mean age of participants was 37.22±13.1 years old. Of the participants, 440 (40.7%) were single, 600 (55.6%) were married and 3.7% were divorced or widower. About 644 (59.7%) were holding a bachelor’s degree, 288 (21.1%) were holding a high school degree or below, and 180 (16.6%) were holding postgraduate degree. Around 508 (47.1%) were employed, 232 (21.5%) were unemployed and 304 (28.2%) were students. Most of participants, 856 (79.3%) were non-Emirati and 224 (20.7%) were Emirati. Many of the participants (52.9%) lived in Sharjah, 288(26.7%) lived in Dubai and the rest 220 (20.3%) lived in the other Emirates.

**Table 1:**
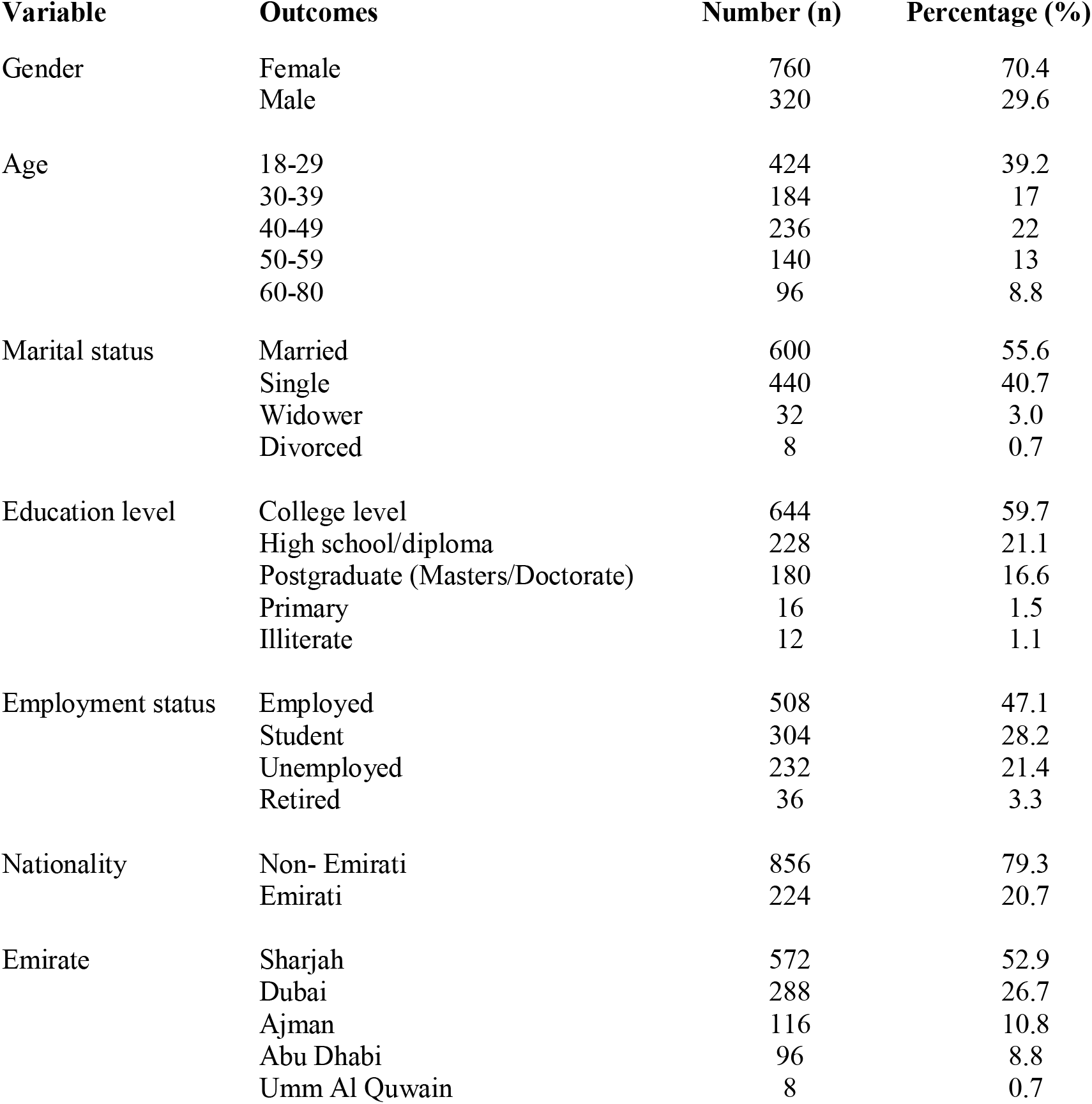
Demographic characteristics of participants, n (1080)

| Variable | Outcomes | Number (n) | Percentage (%) |
| --- | --- | --- | --- |
| Gender | Female | 760 | 70.4 |
|  | Male | 320 | 29.6 |
| Age | 18-29 | 424 | 39.2 |
|  | 30-39 | 184 | 17 |
|  | 40-49 | 236 | 22 |
|  | 50-59 | 140 | 13 |
|  | 60-80 | 96 | 8.8 |
| Marital status | Married | 600 | 55.6 |
|  | Single | 440 | 40.7 |
|  | Widower | 32 | 3.0 |
|  | Divorced | 8 | 0.7 |
| Education level | College level | 644 | 59.7 |
|  | High school/diploma | 228 | 21.1 |
|  | Postgraduate (Masters/Doctorate) | 180 | 16.6 |
|  | Primary | 16 | 1.5 |
|  | Illiterate | 12 | 1.1 |
| Employment status | Employed | 508 | 47.1 |
|  | Student | 304 | 28.2 |
|  | Unemployed | 232 | 21.4 |
|  | Retired | 36 | 3.3 |
| Nationality | Non- Emirati | 856 | 79.3 |
|  | Emirati | 224 | 20.7 |
| Emirate | Sharjah | 572 | 52.9 |
|  | Dubai | 288 | 26.7 |
|  | Ajman | 116 | 10.8 |
|  | Abu Dhabi | 96 | 8.8 |
|  | Umm Al Quwain | 8 | 0.7 |

### Chronic conditions among the participants

Of 1080 participants, 780 (72.2%) were healthy, while 300 (27.8%) had chronic conditions. The most prevalent chronic conditions as shown in **table 2** were diabetes 7.8% followed by hypertension 6.3%, while 3.7%, 3.7%, 3.3%,0.7%, 0.7%, 0.4%, 0.4%, 0.4%, 0.4% suffer of chronic respiratory diseases, heart diseases, obesity, acute cancer, severe anemia, autoimmune diseases, severe allergic, receiving immunotherapy, and liver diseases, respectively.

**Table 2:** Health status and chronic conditions of participants, n (1080)

| Conditions | Number (n) | Percentage (%) |
| --- | --- | --- |
| Acute cancer | 8 | 0.7 |
| Autoimmune diseases | 4 | 0.4 |
| Chronic respiratory diseases | 40 | 3.7 |
| Diabetes | 84 | 7.8 |
| Hypertension | 68 | 6.3 |
| Obesity | 36 | 3.3 |
| Heart disease | 40 | 3.7 |
| Severe allergic reactions to vaccination | 4 | 0.4 |
| Receiving immunotherapy or inhibitor therapy | 4 | 0.4 |
| Severe anemia | 8 | 0.7 |
| Liver diseases | 4 | 0.4 |
| No chronic condition | 780 | 72.2 |

### Relation between the side effects of vaccination with the age (≤49 years old and >49 years old)

Table 3 compares COVID-19-related anamnesis of vaccinated individuals who are below and above the age 49. Regarding whom is vaccinated, the study shows a significant difference between people who are ≤49 years old and >49 years old (p=0.000). Participants who are older than 49 years were more likely to be vaccinated (85%) compared to the ones below 49 years (75%). The results regarding number of doses were not significant (p=0.128). There was a significant difference in whether the participants got Covid-19 infection previously or not with a P value of 0.002. Those who are less or equal to 49 years were more likely to have a previous COVID-19 infection (16.6%).

**Table 3.** COVID-19-related anamnesis of vaccinated individuals, n (1080)

| Anamnesis | Outcome | ≤49 Years Old<br>(844) | >49 Years<br>Old (236) | Total | p-value |
| --- | --- | --- | --- | --- | --- |
| Vaccinated | Yes | 632 (75%) | 200 (85%) | 832 (77%) | 0.000 |
|  | No | 212 (25%) | 36 (15%) | 248(23%) |  |
| Number of doses | One dose | 128 (15%) | 56 (23%) | 184 (17%) | 0.128 |
|  | Two doses | 508 (60%) | 144 (61%) | 652 (60 %) |  |
|  | None | 208 (25%) | 36 (15%) | 244 (23%) |  |
| COVID-19<br>Infection | Yes | 140 (16.6%) | 36 (15%) | 176 (16.1%) | 0.002 |
|  | No | 684 (81%) | 200 (84%) | 884 (81.5%) |  |
|  | Not sure | 20 (2.4%) | 0 | 20 (2.4%) |  |

Table 4. presents the prevalence of general adverse reaction of 1^st^ dose of Sinopharm COVID-19 vaccine in participants (≤49 years old vs >49 years old). The table shows that the around 24.4% (26% vs 18.6%) didn’t get any side effects. The common side effects among both ages of participants were normal pain at the site of vaccination 42.2%, fatigue 12.2%, and headache 9.6%), the same table indicated that was a significant difference between both groups (≤49 years vs >49 years old) with the severe pain at the vaccination site (P= 0.023), nausea (P=0.010) and muscle pain (P=0.010). Moreover, almost 6.8% of participants >49 years reported sever pain at the vaccination site while, compared with 1.42% among ≤49 years old. More proportion of participants who are >49 years old reported nausea and muscle pain compared to patients ≤49 years, it was (0.5% vs 5%) and (4.3% vs 13.6%), respectively.

**Table 4:** Prevalence of the general side effects of after 1^st^ dose Sinopharm COVID-19 vaccination among 2 group of participants (≤49 years old vs >49 years old), no. (1080)

| Side Effect | ≤49 years Old<br>(844), n (%) | >49 years<br>Old (236), n<br>(%) | Total<br>(1080), n (%) | p-value |
| --- | --- | --- | --- | --- |
| Normal pain at the vaccination site | 356 (42.2) | 100 (42.4) | 456 (42.2) | 0.999 |
| Sever pain at the vaccination site | 12 (1.4) | 16 (6.8) | 28 (2.6) | **0.023** |
| Tenderness | 44 (5.2) | 12 (5) | 56 (5.1) | 0.963 |
| Redness | 8 (0.9) | 0 | 8 (0.7) | 0.452 |
| Induration and pruritus at the vaccination site | 12 (1.4) | 0 | 12 (1.1) | 0.356 |
| Fever | 8 (0.9) | 4 (1.7) | 12 (1.1) | 0.631 |
| Headache | 92 (10.9) | 12 (5) | 104 (9.6) | 0.178 |
| Fatigue | 96 (11.4) | 36 (15.3) | 132 (12.2) | 0.429 |
| Nausea | 4 (0.5) | 12(5) | 16 (1.5) | **0.010** |
| Diarrhea | 8 (0.9) | 0 | 8 (0.74) | 0.452 |
| Cough | 8 (0.9) | 4 (1.7) | 12 (1.1) | 0.631 |
| Allergy | 8 (0.9) | 4 (1.7) | 12(1.1) | 0.631 |
| Muscle pain | 36 (4.3) | 32 (13.6) | 68 (6.3) | **0.010** |
| Abdominal Pain | 12 (1.4) | 8 (3.4) | 20 (1.85) | 0.324 |
| Back pain | 24 (2.8) | 20 (8.5) | 44 (4.07) | 0.054 |
| Lethargy | 76 (9) | 24 (10) | 100 (9.2) | 0.631 |
| Others | 12 (1.4) | 0 | 8 (0.7) | 0.452 |
| I did not get any side effect | 220 (26.0) | 44 (18.6) | 264 (24.4) | 0.234 |

The tables shows that no significant between age of participant and another side effects such as normal pain at the vaccination site (P=0.999), tenderness (p=0.963), redness (p=0.452), induration and pruritus at the vaccination site (p=0.356), Fever (p=0.631), headache (p=0.178), fatigue (p=0.429), cough and allergy (p=0.631), Abdominal pain (p=0.324), abdominal pain (p=0.324), back pain (p=0.054), lethargy (p=0.631), and others symptoms (0.452).

In the 2^nd^ dose of Sinopharm COVID-19 vaccine (≤49 years vs >49 years old) there were more prevalence of side effects than the 1^st^ dose (**table 5**). Among the (≤49 years vs >49 years old) participants, 14% (15% vs 10%) didn’t get any post-vaccination symptoms. Furthermore, pain 32.6%, at the vaccination site, fatigue16.3%, lethargy13.7%, headache10%, and tenderness 10% are the most prevalence side effects of the respondents in both groups. The same table shows there was a significant difference between both groups (≤49 years vs >49 years old) and the fatigue (P= 0.003), while no significant difference reported with other side effects.

**Table 5:** Prevalence of the general side effects after 2^nd^ dose Sinopharm COVID-19 vaccination among 2 group of participants (≤49 years old vs >49 years old), no. (1080)

| Side Effect | $\leq 49$ years Old<br>(844), n (%) | $>49$ years Old<br>(236), n (%) | Total<br>(1080), n (%) | p-value |
| --- | --- | --- | --- | --- |
| Normal Pain at the vaccination site | 272 (32.2) | 80 (33.8) | 352 (32.6) | 0.646 |
| Sever pain at the vaccination site | 56 (6.6) | 32 (13.6) | 88 (8.15) | 0.088 |
| Tenderness | 72 (8.5) | 36 (15.3) | 108 (10) | 0.131 |
| Redness | 16 (2) | 0 | 16 (1.5) | 0.285 |
| Induration and pruritus at the vaccination site | 8 (0.95) | 4 (1.7) | 12 (1.1) | 0.631 |
| Fever | 16 (2) | 16 (6.8) | 32 (3) | .051 |
| Headache | 92 (11) | 16 (6.8) | 108 (10) | 0.346 |
| Fatigue | 108 (13) | 68 (28.8) | 176 (16.3) | **0.003\*** |
| Nausea | 8 (0.95) | 4 (1.7) | 12 (1.1) | 0.631 |
| Diarrhea | 8 (0.95) | 0 | 8 (0.7) | 0.452 |
| Cough | 4 (0.5) | 4 (1.7) | 8 (0.7) | 0.336 |
| Allergy | 0 | 0 | 0 | - |
| Muscle pain | 40 (4.7) | 24 (10) | 64 (5.9) | .121 |
| Abdominal Pain | 12 (1.4) | 4 (1.7) | 16 (1.5) | 0.881 |
| Back pain | 20 (2.4) | 12 (5.1) | 32 (3) | 0.28 |
| Lethargy | 104 (12.3) | 44 (18.6) | 148 (13.7) | 0.217 |
| Others | 8 (0.95) | 0 | 8 (0.7) | 0.595 |
| I did not get any side effect | 128 (15) | 24 (10) | 152 (14) | 0.323 |

### Relation between the side effects of vaccination with the gender (females and males)

Table 6 shows the prevalence of side effects post 1^st^ dose of Sinopharm COVID-19 vaccination among two groups (females vs males). Our study was included 760 females and 320 males. The female’s significance had more symptoms of 1^st^ vaccination than males (17% of females didn’t have side effects vs 45% males didn’t have side effects), however, significant relation reported between the symptoms such as fatigue (P=0.006) and gender.

**Table 6:**
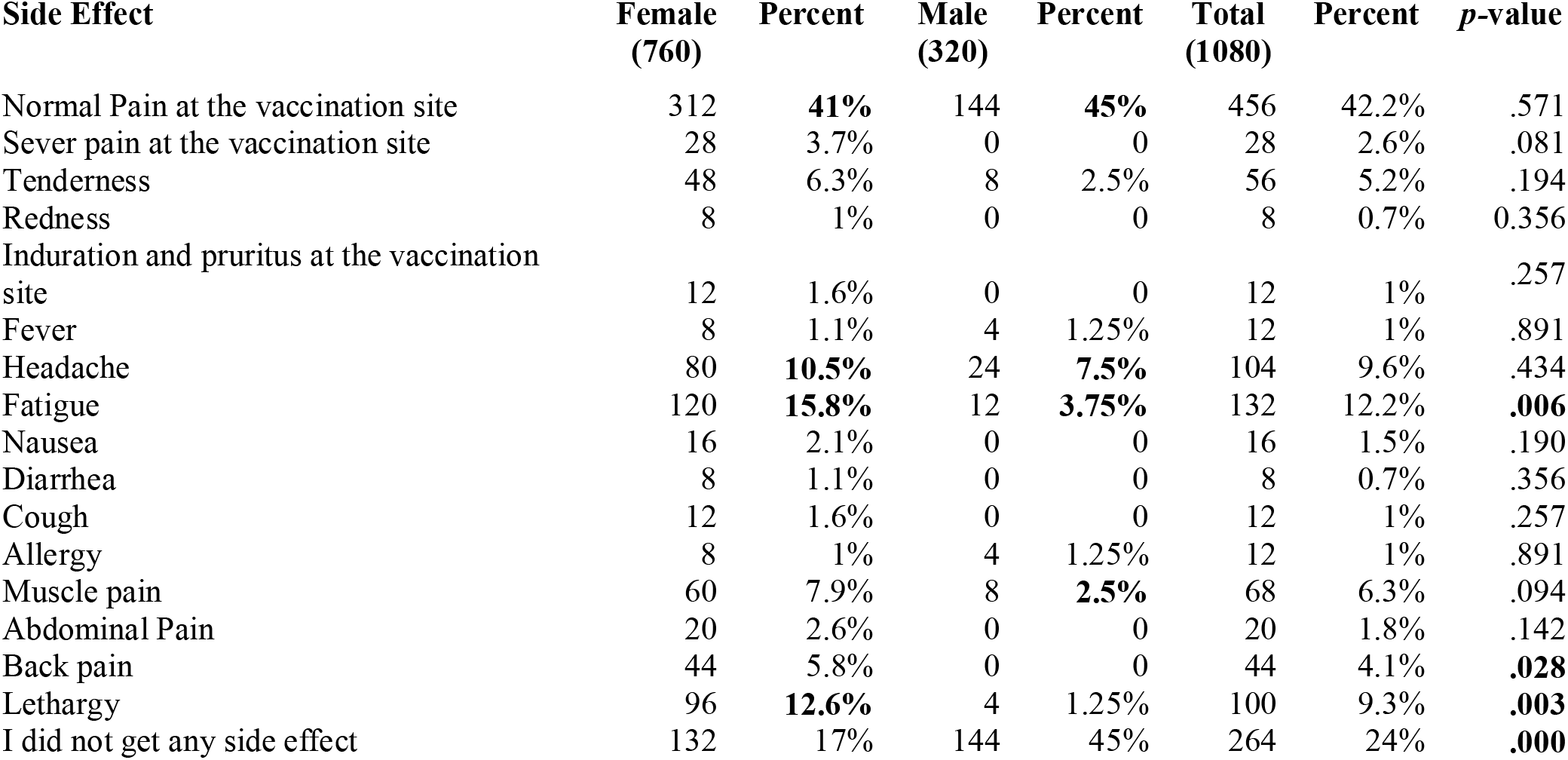
Prevalence of the general side effects after 1^st^ dose Sinopharm COVID-19 Sinopharm COVID-19 vaccination among 2 group of participants (females vs males), n (1080)

The results shows that no significant between the side effects of tenderness (P= 0.194), redness (p-value= 0.356), fever (P= 0.891) and headache (p-value= 0.434) with the gender.

**Table 7** presents the prevalence of side effects of 2nd dose of vaccination among females and males. The females had more side effect of 2^nd^ vaccination dose than males (11.6% of females didn’t have side effects vs 20% males didn’t have side effects), Sever pain at the vaccination site (P=0.027) and fatigue (P = 0.011) symptoms were significantly higher in females than males (10.5% vs 2.5%), (20% vs 7.5%), respectively. In addition, the table shows no significant between the symptoms of normal pain at vaccination site (P=0.482), tenderness (P= 0.368), redness (P = 0.834) fever (P = 0.279), and lethargy (P =0.053) with the genders.

**Table 7:**
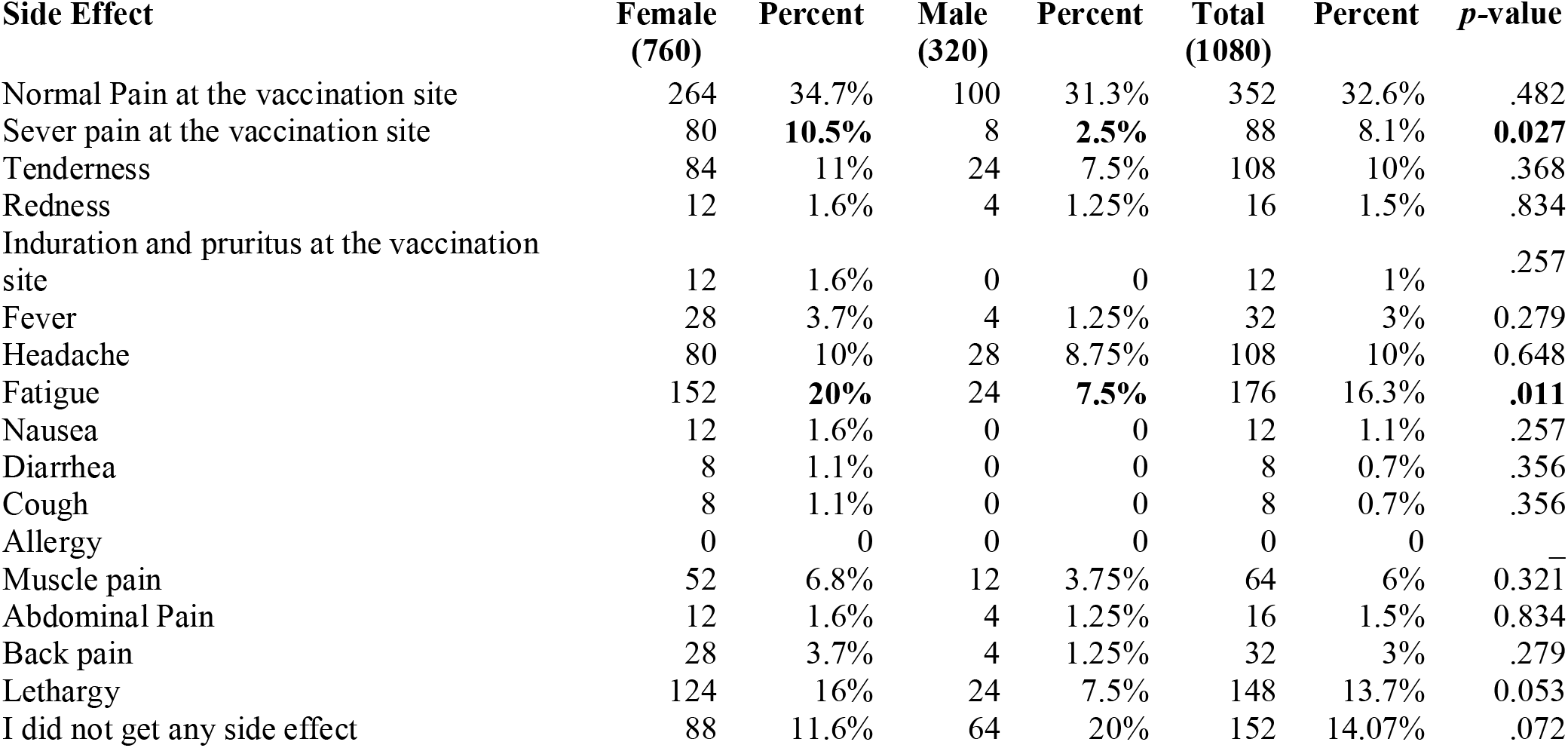
Prevalence of the general side effects after 2^nd^ dose Sinopharm COVID-19 Sinopharm COVID-19 vaccination among 2 group of participants (females vs males), n (1080)

| Side Effect | Female<br>(760) | Percent | Male<br>(320) | Percent | Total<br>(1080) | Percent | p-value |
| --- | --- | --- | --- | --- | --- | --- | --- |
| Normal Pain at the vaccination site | 264 | 34.7% | 100 | 31.3% | 352 | 32.6% | .482 |
| Sever pain at the vaccination site | 80 | **10.5%** | 8 | **2.5%** | 88 | 8.1% | **0.027** |
| Tenderness | 84 | 11% | 24 | 7.5% | 108 | 10% | .368 |
| Redness | 12 | 1.6% | 4 | 1.25% | 16 | 1.5% | .834 |
| Induration and pruritus at the vaccination site | 12 | 1.6% | 0 | 0 | 12 | 1% | .257 |
| Fever | 28 | 3.7% | 4 | 1.25% | 32 | 3% | 0.279 |
| Headache | 80 | 10% | 28 | 8.75% | 108 | 10% | 0.648 |
| Fatigue | 152 | **20%** | 24 | **7.5%** | 176 | 16.3% | **.011** |
| Nausea | 12 | 1.6% | 0 | 0 | 12 | 1.1% | .257 |
| Diarrhea | 8 | 1.1% | 0 | 0 | 8 | 0.7% | .356 |
| Cough | 8 | 1.1% | 0 | 0 | 8 | 0.7% | .356 |
| Allergy | 0 | 0 | 0 | 0 | 0 | 0 | — |
| Muscle pain | 52 | 6.8% | 12 | 3.75% | 64 | 6% | 0.321 |
| Abdominal Pain | 12 | 1.6% | 4 | 1.25% | 16 | 1.5% | 0.834 |
| Back pain | 28 | 3.7% | 4 | 1.25% | 32 | 3% | .279 |
| Lethargy | 124 | 16% | 24 | 7.5% | 148 | 13.7% | 0.053 |
| I did not get any side effect | 88 | 11.6% | 64 | 20% | 152 | 14.07% | .072 |

### Reasons of participants due to don’t take COVID 19 vaccine

The most three common reasons reported by participants for not willing to take the vaccine were 6.3% not believing that the vaccine is effective, 5.2% pant wasn’t authorized to take the vaccine and, 4.4% of them belief that the vaccine have many side effects, Moreover, the participants reported that “not have enough time” 3.7%, unavailability of the vaccine 3%, “the vaccines is not approved by WHO” 1.5%, being afraid of needles 1.1% and the least common reason was “waiting for another vaccine” 0.4% **(table 8)**.

**Table 8:**
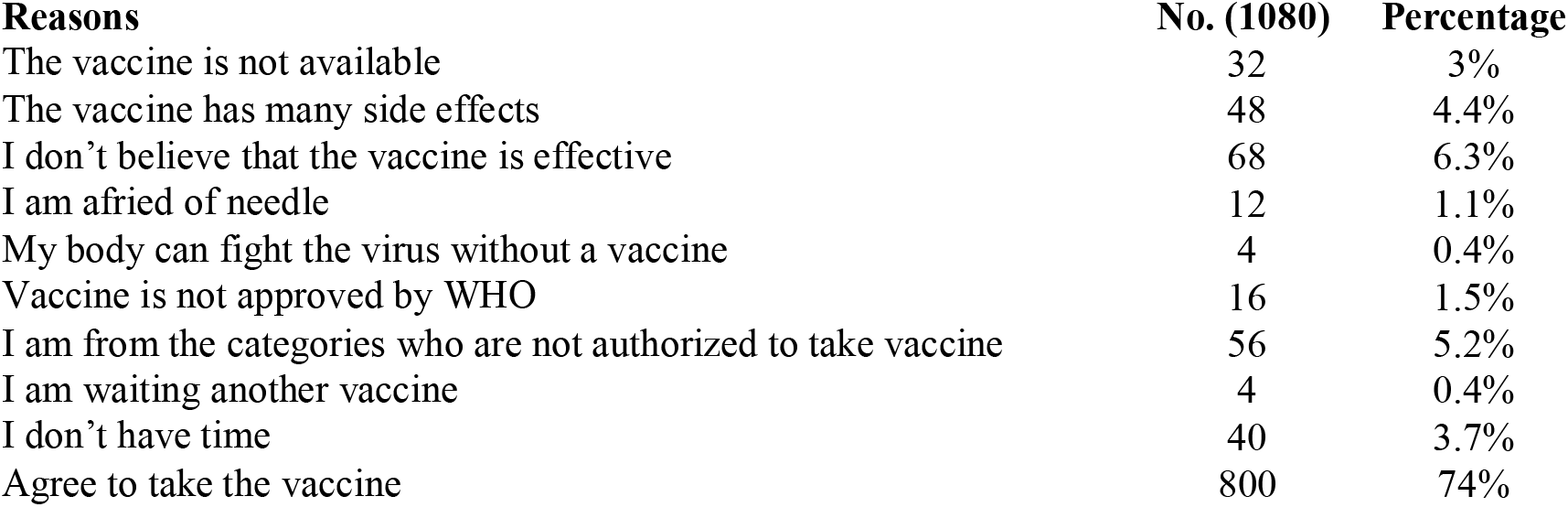
Reasons of participants due to don’t take the COVID 19 vaccine no (%)

## Discussion

in August 2020, Trial 1 and 2 of Sinopharm vaccine were completed and showed that the vaccine triggered a COVID-19 neutralizing antibody response with a low rate of adverse reactions. The most common adverse reactions were pain at the site of injection and fever, but all these were mild and self-limiting, moreover, no treatment was required for any side effect [8]. UAE were among the first to conduct phase three clinical trials of the vaccine, they found that the vaccine to have an 86% efficacy rate according to interim results of its phase 3 trial [8].

Most of studies have assessed mainly the post-vaccination adverse reactions of the Pfizer– BioNTech, Moderna, and Astrazeneca vaccines [21, 22, 23, 24, 25], while only two studies focus on Sinopharm COVID-19 vaccine [16, 17], There are no published studies yet that have focused on Sinopharm COVID-19 vaccine side effects in UAE.

The finding of our study shows that the side effects of this vaccines appear to be mild. Anecdotally, a quarter of participants reported they didn’t get did not get any symptom in post-first vaccination shot, while had mild symptoms following vaccination. Second dose the 14% didn’t report any symptoms, however, most of them had mild and predictable side effects. None of the side effects were of a serious nature or required hospitalization. Our results were in line of a study in India, as frequency of experiencing symptoms following each vaccine were 24.4% (Sinopharm). [16].

In first dose of vaccination the findings showed a statistically significant differences in the prevalence of severe pain at the vaccination site, nausea, and muscle pain, between the participants in both groups (≤49 vs >49). While in the 2nd dose showed a significant difference in the prevalence of Fatigue between two groups. Moreover, the more frequent side effect prevalence is normal Pain at the vaccination site (42.2%), fatigue (12.2%) headache (9.6%), lethargy (9.2%), and muscle pain (6.3%). The reported symptoms in our study were similar to symptoms that appear on participants in phase 1/2 trial of Sinopharm vaccine and reported by Sinopharm vaccine company, WHO, DOH and MOH in the UAE reported that the most adverse reactions were injection site reactions, headaches, fatigue, they indicated that the most common systematic adverse reaction was pain at the vaccination site fever, it was self-limited and recovered, none of the symptoms were of serious nature or requiring hospitalization. [8, 15, 19]

Injection site pain symptom reported in several literatures on vaccines’ side effects [16 Jayadevan, 17, 24, 25], injection in a relaxed muscle leads to less pain compared to a tensed one; therefore, the researchers recommended to lower the patient’s arm that will be injected to reduce the pain. In addition, vaccines in in situ should be keep it in a low temperature, including Sinopharm vaccine that should store at normal refrigeration temperature, and if injected without optimal warming up, this may increase the probability of pain of the injection site symptom. [17, 24, 26].

There was a clear linear correlation between age and vaccination side effects, the younger adults (≤49) were more frequently affected than other groups (>49). Vaccine reactogenicity is known to correlate with transient elevation of inflammatory cytokines, suggesting that the vaccine reactogenicity declined with age. but it is not considered a reliable sign of a desirable immune response [27]. Similar to our results showed in various studies on COVID-19 vaccine side effects [16, 24, 28], Moreover, according to the Centers for Disease Control and Prevention (CDC), side effects tend to be more noticeable after the first dose [29].

Similar to findings of studies recently published [17, 25] We observed that the frequency of adverse effective in the second shot was slightly higher than to the first dose. Except some symptoms (1st vs 2nd), nausea (1.5% vs 1.1%), Allergy (1.1% vs 0), Cough (1.1% vs 0.7%), Abdominal Pain (1.85% vs 1.5%, Back pain (4.07% vs 3%), this finding could be interpreted on the basis response of the immune system’s. The immune system could produce cytokines that could have an inflammatory effect on the blood vessels, muscles, and other tissues. It may also produce flu-like symptoms that last for days after vaccination. [7].

Women generally were more likely to develop side effects of vaccination than males, as 83% of the females reported they had side effects compared to 55% reported by males’ participants post 1st dose of vaccination, while was (98.5% female vs, 80% males) for post 2nd dose of vaccination. Previous studies on COVID-19 vaccine have reported more side effect in females compared to males in both doses [16, 24, 25].

According to our results the (1.1%) of participants has been Allergic symptoms post -1st vaccination, while no cases for allergy reported among them post -2nd dose vaccination.

This confirms that the participants with allergic reaction in this study did not receive the second dose of vaccination, there are no published data on the safety of the 2nd dose of covid vaccine after an allergic reaction to the first dose. one study indicated that anyone with immediate allergic reaction history of any severity to any component of mRNA COVID-19 vaccines or to polyethylene glycol or polysorbate should not be vaccinated with the Pfizer-BioNTech or Moderna COVID-19 vaccine.[30].

Allergic reactions to vaccines not attributed to the active vaccine itself, they might be caused by inactive ingredients, such as egg protein, gelatin, formaldehyde, thimerosal, or neomycin, that contribute to specific IgE-mediated immediate reactions. According to European Medicines Agency (EMA), Excipients, are constituents of a medicinal form apart from the active substance, it constitute inert substances that are added to vaccines to improve stability, increase solubility, improve absorption, influence palatability, or create a distinctive appearance. Excipients can cause various clinical allergic reactions ranging from skin disorders to life-threatening systemic reactions [31].

In our study, we noticed that the most prevalent chronic conditions among the UAE participants were diabetes 7.80%, and hypertension 6.30%, our results are consistent with the Dubai Statistical Center study that shows UAE nationals have a higher prevalence rate of diabetes and Hypertension diseases, furthermore, another study among patients with chronic disease in the UAE reported that 74.1% were Diabetics [32, 33].

Recent studies have shown that COVID-19 vaccine hesitancy level among residents varies from low to high, 29% of New York residents claiming they will refuse a vaccine, compared to 20% of Canadians residents, and 6% of among them in the United Kingdom. [34, 35].

In our study, a few percentages of the respondents reported that don’t want to receive the COVID-19 vaccine, the most reasons of indecision and rejection about COVID-19 were belief that the vaccine not effective, some of the participants were from the categories who are not authorized to take vaccine, while 4.4% were afraid of side effects of the vaccine. Several studies in the scientific literature reported the similar reasons, Thus, knowing what to expect post-vaccination from this study will help with public education, dispelling myths, and lowering the apprehension about Sinopharm COVID-19 vaccines.

similar reasons reported in several studies, as the side effects of vaccine, don’t think the vaccine can be reliable as it will be a new vaccine, and COVID-19 infection is a biological weapon and the vaccine will serve those who produce this virus, were the most common reasons for rejection of vaccine [36]

## Conclusion

Fear of the unknown is a driver of vaccine hesitancy. This study showed that 1st and 2nd dose post-vaccination adverse reactions of Sinopharm COVID-19 vaccine were common side effects, mild, predictable, non-life-threatening, and none were serious. To our knowledge, this is the first design dealing with the Sinopharm vaccine side and evaluating the side effects among a UAE population, the results may help reduce vaccine hesitancy of the public.

## Data Availability

all data available

## References

1. Habas, K., Nganwuchu, C., Shahzad, F., Gopalan, R., Haque, M., & Rahman, S. et al. (2020). Resolution of coronavirus disease 2019 (COVıD-19). Expert Review Of Anti-ınfective Therapy, 18(12), 1201–1211. doi: 10.1080/14787210.2020.1797487

2. Lai C, Shih T, Ko W, Tang H, Hsueh P. Severe acute respiratory syndrome coronavirus 2 (SARS-CoV-2) and coronavirus disease-2019 (COVıD-19): The epidemic and the challenges. Agents. 2020; 55(3):105924

3. Coronavirus disease (COVıD-19): How is it transmitted?. (2021). Retrieved 1 June 2021, from https://www.who.int/emergencies/diseases/novel-coronavirus-2019/question-and-answers-hub/q-a-detail/coronavirus-disease-covid-19-how-is-it-transmitted

4. Investigational treatments for COVıD-19. (2020). The Pharmaceutical Journal. doi: 10.1211/pj.2020.20208051

5. Haidere, M., Ratan, Z., Nowroz, S., Zaman, S., Jung, Y., Hosseinzadeh, H., & Cho, J. (2021). COVıD-19 Vaccine: Critical Questions with Complicated Answers. Biomolecules & Therapeutics, 29(1), 1–10. doi: 10.4062/biomolther.2020.178

6. Petousis-Harris, H. (2021). Correction to: Assessing the Safety of COVıD-19 Vaccines: A Primer. Drug Safety, 44(4), 507–507. doi: 10.1007/s40264-020-01023-1

7. Zhang, Y., Zeng, G., Pan, H., Li, C., Hu, Y., & Chu, K. et al. (2021). Safety, tolerability, and immunogenicity of an inactivated SARS-CoV-2 vaccine in healthy adults aged 18–59 years: a randomised, double-blind, placebo-controlled, phase 1/2 clinical trial. The Lancet ınfectious Diseases, 21(2), 181–192. doi: 10.1016/s1473-3099(20)30843-4

8. Xia, S., Duan, K., Zhang, Y., Zhao, D., Zhang, H., Xie, Z., … Yang, X. (2020). Effect of an inactivated vaccine against SARS-CoV-2 on safety and immunogenicity outcomes: ınterim analysis of 2 randomized clinical trials. JAMA, 324(10), 951–960. doi:10.1001/jama.2020.15543

9. Forni, G.; Mantovani, A. COVıD-19 Commission of Accademia Nazionale deiLincei, Rome. COVıD-19 vaccines: Where we stand and challenges ahead. Cell Death Differ. 2021, 28, 626–639.

10. Sharma O, Sultan AA, Ding H, Chris R. A Review of the Progress and Challenges of Developing a Vaccine for COVıD-19. Front. ımmunol; 2020; 11,. 585354.

11. China to run human coronavirus vaccine trial in UAE. Reuters 2020 Jun 23. https://www.reuters.com/article/us-health-coronavirus-china-vaccine/china-to-run-human-coronavirus-vaccine-trial-in-uae-idUSKBN23U2H8.

12. Development of an ınactivated Vaccine Candidate, BBıBP-CorV, with Potent Protection against SARS-CoV-2. (2021). Retrieved 1 June 2021, from https://dx.doi.org/10.1016%2Fj.cell.2020.06.008

13. Cyranoski, D. (2020). Arab nations first to approve Chinese COVıD vaccine — despite lack of public data. Nature, 588(7839), 548. https://doi.org/10.1038/d41586-020-03563-z

14. Zahid MN, Moosa MS, Perna S, Ebtisam EB. A review on COVıD-19 vaccines: stages of clinical trials, mode of actions and efficacy. Arab Journal of Basic and Applied Sciences.2021; 28:1, 225–233.

15. World Health Organization (WHO): Evidence Assessment: Sinopharm/BBıBP COVıD-19 vaccine, for recommendation by the strategic advisory group of experts (sage) on immunization prepared by the sage working group on covid-19 vaccines. 2021

16. Jayadevan R, Shenoy R, Anithadevi TS. Survey of symptoms following COVıD-19 vaccination in ındia.medRxiv preprint doi: https://doi.org/10.1101/2021.02.08.21251366.

17. Hatmal MM, Al-Hatamleh MAı, Olaimat AN, Hatmal M, Alhaj-Qasem DM, Olaimat TM, Mohamud R. Side Effects and Perceptions Following COVıD-19 Vaccination in Jordan: A Randomized, Cross-Sectional Study ımplementing Machine Learning for Predicting Severity of Side Effects. Vaccines 2021, 9, 556.

18. Aresté N, Salgueira M. World Medical Association Declaration of Helsinki: ethical principles for medical research involving human subjects. JAMA. 2013;310(20):2191–4.

19. COVıD-19 Vaccine, Awareness Guide, April, 2021, https://www.dha.gov.ae/Asset%20Library/COVID19/Covid19_Vaccine_EN.pdf

20. Nunnally, J.C. and Bernstein, ı.H. (1994) The Assessment of Reliability. Psychometric Theory, 3, 248–292.

21. Kadali, R.A.K.; Janagama, R.; Peruru, S.; Malayala, S.V. Side effects of BNT162b2 mRNA COVıD-19 vaccine: A randomized, cross-sectional study with detailed self-reported symptoms from healthcare workers. ınt. J. ınfect. Dis. 2021, 106, 376–381.

22. Chapin-Bardales, J.; Gee, J.; Myers, T. Reactogenicity Following Receipt of mRNA-Based COVıD-19 Vaccines. JAMA 2021

23. Menni, C.; Klaser, K.; May, A.; Polidori, L.; Capdevila, J.; Louca, P.; Sudre, C.H.; Nguyen, L.H.; Drew, D.A.; Merino, J.; et al. Vaccine side-effects and SARS-CoV-2 infection after vaccination in users of the COVıD Symptom Study app in the UK: A prospective observational study. Lancet ınfect. Dis. 2021.

24. Riad A, Pokorná A, Attia S, Klugarová J, Koščík M, Klugar M. Prevalence of COVıD-19 Vaccine Side Effects among Healthcare Workers in the Czech Republic. 2021. J. Clin. Med. 2021, 10, 1428.

25. El-Shitany NA, Harakeh S, Badr-Eldin SM, Bagher AM, Eid B, Almukadi H, Alghamdi BS, Alahmadi AA, Hassan NA, Sindi N, Alghamdi SA, Almohaimeed HM, Mohammedsaleh ZM, Al-Shaikh TM, Almuhayawi MS, Ali SS, El-Hamamsy M. Minor to Moderate Side Effects of Pfizer-BioNTech COVıD-19 Vaccine Among Saudi Residents: A Retrospective Cross-Sectional Stud. ınternational Journal of General Medicine 2021:14 1389–1401.

26. “China State-Backed Covid Vaccine Has 86% Efficacy, UAE Says “. Bloomberg.com. 2020-12-09. Retrieved 2020-12-09.

27. Hervé, C., Laupèze, B., Del Giudice, G. et al. The how ‘s and what ‘s of vaccine reactogenicity. npj Vaccines 4, 39, 2019 https://doi.org/10.1038/s41541-019-0132-6

28. Polack F, Thomas S, Kitchin N, et al. Safety and efficacy of the BNT162b2 mRNA covid-19 vaccine. N Engl J Med. 2020; 383:2603–2615. doi:10.1056/NEJMoa2034577

29. Possible Side Effects After Getting a COVıD-19 Vaccine. Available online: https://www.cdc.gov/coronavirus/2019-ncov/vaccines/expect/after.html (accessed on 25 April 2021).

30. Kounis NG, Koniari ı, Gregorio CD, Velissaris D, Petalas K, Brinia A, Assimakopoulos SF, Gogos C, Kouni SN, Kounis GN, Calogiuri G, Ming-Yow Hung M. Allergic Reactions to Current Available COVıD-19 Vaccinations: Pathophysiology, Causality, and Therapeutic Considerations. Vaccines 2021, 9(3), 221

31. Caballero, M.L.; Quirce, S. Delayed Hypersensitivity Reactions Caused by Drug Excipients: A Literature Review. J. ınvestig. Allergol. Clin. ımmunol. 2020, 30, 400–408

32. CDC. 2020 https://www.cdc.gov/coronavirus/2019-ncov/need-extra-precautions/people-with-medical-conditions.html

33. Osama H, ıbrahim M, Jirjees FJ, Mahdi HM. Barriers affecting compliance of patients with chronic diseases: A preliminary study in United Arab Emirates (UAE) population. AJPCR 2011; 4: suppl 2.

34. Latimer K. About 20% of people in recent survey said they wouldn ‘t take COVıD-19 vaccine; 2020. https://www.cbc.ca/news/canada/saskatchewan/covid-survey-first-round-results-1.5541053. Accessed July 27, 2020.

35. Henley J; Guardian correspondents. Coronavirus causing some anti-vaxxers to waver, experts say; 2020. https://www.theguardian.com/world/2020/apr/21/anti-vaccination-community-divided-how-respond-to-coronavirus-pandemic. Accessed July 27, 2020.

36. Akarsu B, Özdemir DC, Baser DA, Aksoy H, Fidancı ı, Cankurtaran M. While studies on COVıD-19 vaccine is ongoing, the public ‘s thoughts and attitudes to the future COVıD-19 vaccine. ınt J Clin Pract. 2021;75: e13891.

